# An integrated multi-omics analysis identifies clinically relevant molecular subtypes of non-muscle-invasive bladder cancer

**DOI:** 10.1101/2020.06.19.20054809

**Authors:** Sia Viborg Lindskrog, Frederik F. Prip, Philippe Lamy, Ann Taber, Clarice S. Groeneveld, Karin Birkenkamp-Demtröder, Jørgen Bjerggaard Jensen, Trine Strandgaard, Iver Nordentoft, Emil Christensen, Mateo Sokac, Nicolai J. Birkbak, Lasse Maretty, Gregers G. Hermann, Astrid C. Petersen, Veronika Weyerer, Marc-Oliver Grimm, Marcus Horstmann, Gottfrid Sjödahl, Mattias Höglund, Torben Steiniche, Karin Mogensen, Aurélien de Reyniès, Roman Nawroth, Brian Jordan, Xiaoqi Lin, Dejan Dragicevic, Douglas G. Ward, Anshita Goel, Carolyn D. Hurst, Jay D. Raman, Joshua I. Warrick, Ulrika Segersten, Danijel Sikic, Kim E.M. van Kessel, Tobias Maurer, Joshua J. Meeks, David J. DeGraff, Richard T. Bryan, Margaret A. Knowles, Tatjana Simic, Arndt Hartmann, Ellen C. Zwarthoff, Per-Uno Malmström, Núria Malats, Francisco X. Real, Lars Dyrskjøt

**Author notes:** Equal contribution (Sia Viborg Lindskrog, Frederik F. Prip).

## Abstract

The molecular landscape in non-muscle-invasive bladder cancer (NMIBC) is characterized by large biological heterogeneity with variable clinical outcomes. Here, we performed a large integrative multi-omics analysis of patients diagnosed with NMIBC (n=834). Transcriptomic analysis identified four classes (1, 2a, 2b and 3) reflecting tumor biology and disease aggressiveness. Both transcriptome-based subtyping and the level of chromosomal instability provided independent prognostic value beyond established prognostic clinicopathological parameters. High chromosomal instability, p53-pathway disruption and APOBEC-related mutations were significantly associated with transcriptomic class 2a and poor outcome. RNA-derived immune cell infiltration was associated with chromosomally unstable tumors and enriched in class 2b. Spatial proteomics analysis confirmed the higher infiltration of class 2b tumors and demonstrated an association between higher immune cell infiltration and lower recurrence rates. Finally, a single-sample classification tool was built and the independent prognostic value of the transcriptomic classes was documented in 1306 validation samples. The classifier provides a framework for novel biomarker discovery and for optimizing treatment and surveillance in next-generation clinical trials.

## Introduction

Urothelial non-muscle-invasive bladder cancer (NMIBC) represents the most common type of bladder cancer. Patients with NMIBC experience a high likelihood of disease recurrence (50-70%) and progression to muscle-invasive bladder cancer (MIBC; up to 20%, depending on stage and grade) ^1^. Consequently, although 5-year survival rates are favorable (>90%), most patients must undergo lifelong cystoscopic surveillance and multiple therapeutic interventions, making bladder cancer the most expensive cancer to treat ^2^. Clinically, high-risk NMIBC is treated with adjuvant intravesical instillations of Bacillus Calmette–Guérin (BCG) after surgery to eradicate residual disease and hence reduce the frequency of recurrence and progression ^1^.

Despite similar clinical and histopathological characteristics, tumors show large differences in disease aggressiveness and response to therapy, emphasizing the urgent need for further delineation of clinically useful biomarker tests to facilitate and improve patient surveillance and treatment ^3^. Earlier studies of NMIBC biology addressed gene expression for classification of aggressiveness, resulting in the identification of two major molecular subtypes ^4–6^. Thereafter, five subtypes of bladder cancer were identified when the whole spectrum of disease stages was considered; in particular, three subtypes (Urothelial-like, Genomically Unstable, and a group of infiltrated cases) were associated with NMIBC ^7^. In a more recent study of 460 NMIBC patients, we reported three gene expression-based classes (class 1-3; UROMOL2016 classification system) with different clinical outcomes and molecular characteristics ^8^. Large differences in biological processes, such as cell cycle, epithelial-mesenchymal transition (EMT) and differentiation, were observed. Furthermore, mutations in well-known cancer driver genes, i.e *TP53* and *ERBB2*, were primarily found in high-risk class 2 tumors, together with enrichment for APOBEC-related mutational processes.

Analysis of genomic alterations in NMIBC has revealed complex genomic patterns underlying bladder carcinogenesis. Activating mutations in *FGFR3* and *PIK3CA* and chromosome 9 deletions have been identified as early disease drivers ^9–11^. Recently, van Kessel et al. showed that NMIBC at high risk for progression could be further subdivided into good, moderate and poor progression risk groups based on mutations in *FGFR3* and methylation of *GATA2* ^12^. Hurst et al. assessed 160 tumors for genome-wide copy number alterations (CNAs) using array-based comparative genomic hybridization (CGH). The study included 49 high-grade T1 tumors which separated into three major genomic subgroups, one of which contained the majority of tumors showing disease progression ^13^. In a more recent study, the same group analyzed CNAs in 140 Ta tumors and identified two major genomic subtypes (GS1 and GS2). GS1 tumors showed no or very few CNAs, while tumors in GS2 showed more alterations and a high frequency of chromosome 9 deletions ^14^. Exome sequencing of 28 Ta tumors revealed that GS2 tumors had a higher mutational load with enrichment for APOBEC-related mutations compared to GS1 tumors. Furthermore, comparing 79 of the samples to transcriptional subtypes showed that the tumors were primarily classified as the Urothelial-like A subtype (Lund Taxonomy). Application of the UROMOL2016 classification system showed that GS2 tumors with higher genomic instability were enriched for the class 2 subtype ^14^. However, additional refinement of these genomic studies is required to determine optimal predictors of disease aggressiveness and outcome.

The tumor microenvironment has also been linked to prognosis in NMIBC. A high infiltration of cytotoxic T lymphocytes (CTLs) is associated with better prognosis in many cancer types, including MIBC ^15,16^. In contrast, high infiltration of tumor infiltrating-lymphocytes (TILs) has been associated with progression in NMIBC ^17,18^. Furthermore, the presence of tumor-associated macrophages and mature tumor-infiltrating dendritic cells has been related to progression of NMIBC ^19^. The impact of regulatory T cells (Tregs) is conflicting, since high infiltration of Tregs has been associated with both a favorable ^20^ and unfavorable prognosis of bladder cancer ^21,22^. The impact of immune cell infiltration on disease outcome and association with molecular subtypes and genomic alterations in NMIBC needs to be further studied.

Overall, our understanding of the molecular landscape of NMIBC is still incomplete, and integrative multi-omics layered analysis is needed to obtain further knowledge of biological processes contributing to disease aggressiveness, recurrence and progression. This should ultimately lead to biomarker-based optimized surveillance and therapy modalities for patients with NMIBC. Here, we report the largest integrative multi-omics analysis of NMIBC tumors from a total of 834 patients included in the UROMOL project. With this analysis, we delineate genomic and transcriptomic predictors of outcome in NMIBC, and present an online tool for classification of independent samples (**http://cit.ligue-cancer.net:3838/apps/BLCAclassify/**).

## Results

### Clinical, pathological and molecular information

Patients were enrolled in the UROMOL project, a European multicenter prospective study of NMIBC. The initial reports from the UROMOL project included only transcriptomic analysis ^6,8^. We have now performed an integrated multi-omics analysis and have expanded the work to a larger NMIBC patient series with updated follow-up that is essential to acquire insight into the implications for patient management. In total, 862 tumors (611 Ta, 240 T1, 11 carcinoma *in situ* (CIS)) were analyzed in this study. Median follow-up for patients without progression was 49 months and 10.3% progressed to MIBC. A detailed summary of clinical and histopathological information and the analyses performed is provided in **Table S1**.

### Delineation of transcriptomic classes in NMIBC

We analyzed bulk RNA-Seq data from 535 patients (395 Ta, 137 T1, 3 CIS; an expansion of the 460 NMIBC patient cohort previously analyzed ^8^). Using unsupervised consensus clustering of gene-based expression values restricted to the 4000 genes with highest variation across the dataset ^23^, we identified four transcriptomic classes which partially overlapped with the previous UROMOL2016 classes 1-3: high-risk class 2 was further subdivided into two subclasses, named class 2a and 2b for continuity (**Fig 1A**). Classes showed significantly different progression-free survival (PFS, p=6.6 × 10^−5^; **Fig 1B**): patients with class 2a tumors had the worst outcome, followed by patients with class 2b tumors. Patients with class 1 tumors had the best recurrence-free survival (RFS, p=0.025; **Fig 1C**). Multivariable Cox regression analysis revealed that high-risk class 2a and 2b were independently associated with worse PFS and RFS when adjusted for the clinical EORTC (European Organisation for Research and Treatment of Cancer, ^24^) risk score (**Table S2**).

**Figure 1.**
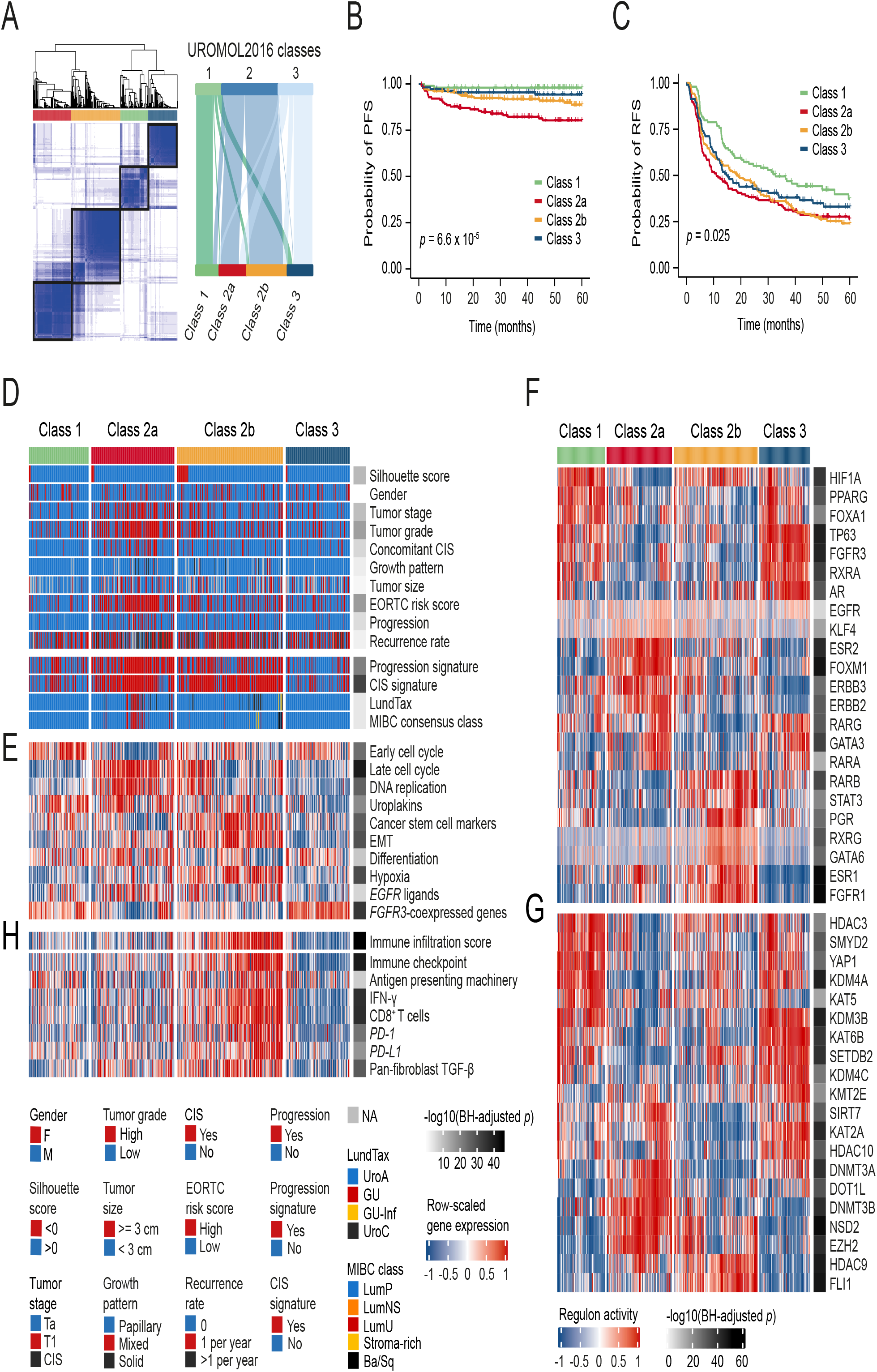
Transcriptomic classes in NMIBC. **a Left:** consensus matrix for four clusters. Samples are in both rows and columns and pairwise values range from 0 (samples never cluster together; white) to 1 (samples always cluster together; dark blue). **Right**: comparison between the three UROMOL2016 transcriptomic classes and the UROMOL2020 four-cluster solution (76% of tumors in UROMOL2016 class 1 remained class 1, 92% of tumors in UROMOL2016 class 2 remained class 2a/2b and 67% of tumors in UROMOL2016 class 3 remained class 3). **b** Kaplan-Meier plot of PFS for 530 patients stratified by transcriptomic class. **c** Kaplan-Meier plot of RFS for 511 patients stratified by transcriptomic class. **d-e** Clinicopathological information and selected gene expression signatures for all patients stratified by transcriptomic class. Samples are ordered after increasing silhouette score within each class (lowest to highest class correlation). **f** Regulon activity profiles for 23 transcription factors. Samples are ordered as in (D). Regulons are hierarchically clustered. **g** Regulon activity profiles for potential regulators associated with chromatin remodeling. The most-upregulated regulons within each class are shown. Regulons are hierarchically clustered. **h** RNA-based immune score and immune-related gene expression signatures for all patients stratified by transcriptomic class.

Transcriptomic classes were significantly associated with various clinicopathological parameters (**Fig 1D** and **Table S3**). Class 2a was enriched for T1 tumors, high-grade tumors, and tumors from patients with CIS and high EORTC risk scores (**Table S3**). Tumors in class 2a also showed a significant overlap with those expressing our previously reported progression signature (80%; p=5.5 × 10^−21^; ^4,6^). Similarly, class 2a and 2b tumors strongly overlapped with those having a positive CIS signature (77% and 87% respectively; p=4.3 × 10^−32^; ^5^). Classification using the Lund system ^25^ revealed that 91% of tumors were classified as UroA and 4% as genomically unstable (GU), the latter mostly found in class 2a (**Fig 1D**). When classified according to the six consensus classes of MIBC ^26^, 93% of tumors classified as Luminal Papillary (LumP). Consequently, the new classification provided here captures the molecular granularity of NMIBC to an extent that previous strategies were unable to.

Analysis of the biological processes associated with NMIBC classes revealed important information discriminating classical histological features from molecular classification and outcome. Confirming our previous findings, class 1 and 3 tumors were associated with early cell cycle genes (p=1.1 × 10^−15^ and p=0.003, respectively; **Fig 1E**). By contrast, class 2a tumors were most strongly associated with late cell cycle genes (p=1.3 × 10^−33^), DNA replication (p=1.1 × 10^−20^), uroplakins (p=9.1 × 10^−7^) and genes involved in cell differentiation (p=2.9 × 10^−5^), thus indicating that differentiation and proliferation do not show an inverse association. Additionally, class 2b tumors were predominantly associated with expression of cancer stem cell (CSC) markers (p=9.7 × 10^−25^) and genes involved in EMT (p=7.3 × 10^−24^), but a lesser association with cell proliferation (**Fig 1E** and **Fig S1A**).

To explore transcriptomic differences between NMIBC classes further, we analyzed transcriptional regulatory networks (i.e. regulons) for a predefined list of 23 transcription factors previously investigated for MIBC ^27^ and candidate regulators associated with chromatin remodeling in cancer ^28^. This analysis provided strong confirmation of the biological relevance of a four-subtype classification, as regulon activities were highly associated with transcriptomic classes (**Fig 1F-G**). Similar regulon activity patterns were shared by class 1 and 3 tumors, but class 3 tumors differed by having high AR and GATA3 regulon activity. Class 2a tumors were distinctly associated with high FOXM1, ESR2, ERBB2 and ERBB3 regulon activity, while class 2b tumors showed high activity of the ESR1, FGFR1, RARB, STAT3 and PGR regulons. Activity profiles of regulons associated with chromatin remodeling highlighted additional potential regulatory differences between class 1 and 3 tumors, indicating that epigenetic-driven transcriptional networks (e.g. KMT2E, KAT2A, KAT5, HDAC10) might be important differentiators of these classes (**Fig 1G** and **Fig S1B**). The potential epigenetic differences between the classes were further supported by an EPIC BeadChip methylation analysis of 29 Ta high-grade tumors, which demonstrated an overall large difference in promoter methylation between samples from different classes (**Fig S1C**). Furthermore, when comparing class 1 and 3 tumors it was revealed that gene promoters were less methylated in class 3 (**Fig S1D**). In total, 12,035 promoter sites were differentially methylated between class 1 and 3 tumors and of these, 97.9 % were more methylated in class 1 compared to class 3.

We estimated the presence of immune cells by deconvolution of RNA-Seq data ^29^. Class 2b tumors had a significantly higher total immune infiltration score compared to all other classes (p=1.3 × 10^−43^), indicating a high level of immune cell infiltration (**Fig 1H**). Class 3 tumors had a significantly lower immune infiltration score compared to both class 1 and 2a (p=1.8 × 10^−7^). Since class 2b tumors showed a favorable PFS compared to class 2a tumors (p=0.024, **Fig S1E**), we investigated the prognostic impact of immune infiltration irrespective of NMIBC class. The transcriptome-based measure of immune infiltration was, however, not associated with PFS or RFS *per se* (**Fig S1F-G**). We also characterized the four classes using gene signatures of potential relevance for different treatment strategies (**Fig 1E and 1H**). Class 2b tumors showed significantly higher expression of immune checkpoint markers and other immune-related signatures compared to all other classes, suggesting that such tumors might be more responsive to immunotherapies ^30,31^. However, no difference in BCG failure-free survival was observed between patients with high-grade class 2a or 2b tumors treated with a minimum of six BCG cycles (n=54, **Fig S1H**).

### Chromosomal instability is associated with high-risk NMIBC

To investigate the genomic heterogeneity of NMIBC further, a total of 473 tumor-leukocyte pairs were analyzed using Illumina SNP arrays. Genomic losses/gains and allelic imbalance were derived from raw segmented total copy-number and B allele frequency (BAF) values (for details see **Methods**). Analysis of the genomic landscape in tumors stratified by EORTC risk score showed similar patterns of abnormalities, but genomic alterations (except for chromosome 9 losses) were more frequently found in EORTC high-risk tumors (**Fig S2A**). Tumors were therefore stratified to three genomic classes (GC1-3) of equal size with increasing CNA burden to illustrate low, intermediate and high chromosomal instability (**Fig 2A** and **Fig S2B**). The distribution of clinicopathological parameters and molecular variables between the genomic classes is shown in **Fig 2A**. Specifically, we observed partial or complete loss of chromosome 9 in 53% (251/473; *CDKN2A* loci) of tumors, and amplification of 8q22.1 in 22% of tumors (103/473; *GDF6* and *SDC2* loci). Genes in the affected 8q22.1 loci may be involved in the dysregulation of extracellular matrix synthesis and transforming growth factor (TGF)-β pathway ^32^. Other frequently altered genomic areas included gains of 1q (16%), 8q (14%; including *MYC*), 5p (11%; including *TERT*), 20q (11%) and 20p (9.3%), and losses on 8p (16%), 11p (14%), 17p (13%; including *TP53*) and 18q (8.2%). Genomic classes were significantly associated with PFS and RFS (p=1.5 × 10^−7^ and p=1.5 × 10^−5^, respectively; **Fig 2B-C**). Importantly, restricting the survival analysis to tumors with high EORTC risk score (>6), genomic classes were still significantly associated with PFS (**Fig 2D**). Genomic classes were significantly associated with stage, grade, concomitant CIS and EORTC risk score (**Fig 2A** and **Table S4**); however, multivariable Cox regression analysis documented that genomic classes were an independent prognostic variable for progression when adjusted for tumor stage and grade (HR=3.5 (95%CI: 1.57-7.56); p=0.002) and EORTC risk score (HR=2.8 (95%CI: 1.28-5.99); p=0.01) (**Table S2**). Furthermore, genomic classes were also independently associated with recurrence when adjusted for EORTC risk score (HR=1.5 (95%CI: 1.13-2.04); p=0.005).

**Figure 2.**
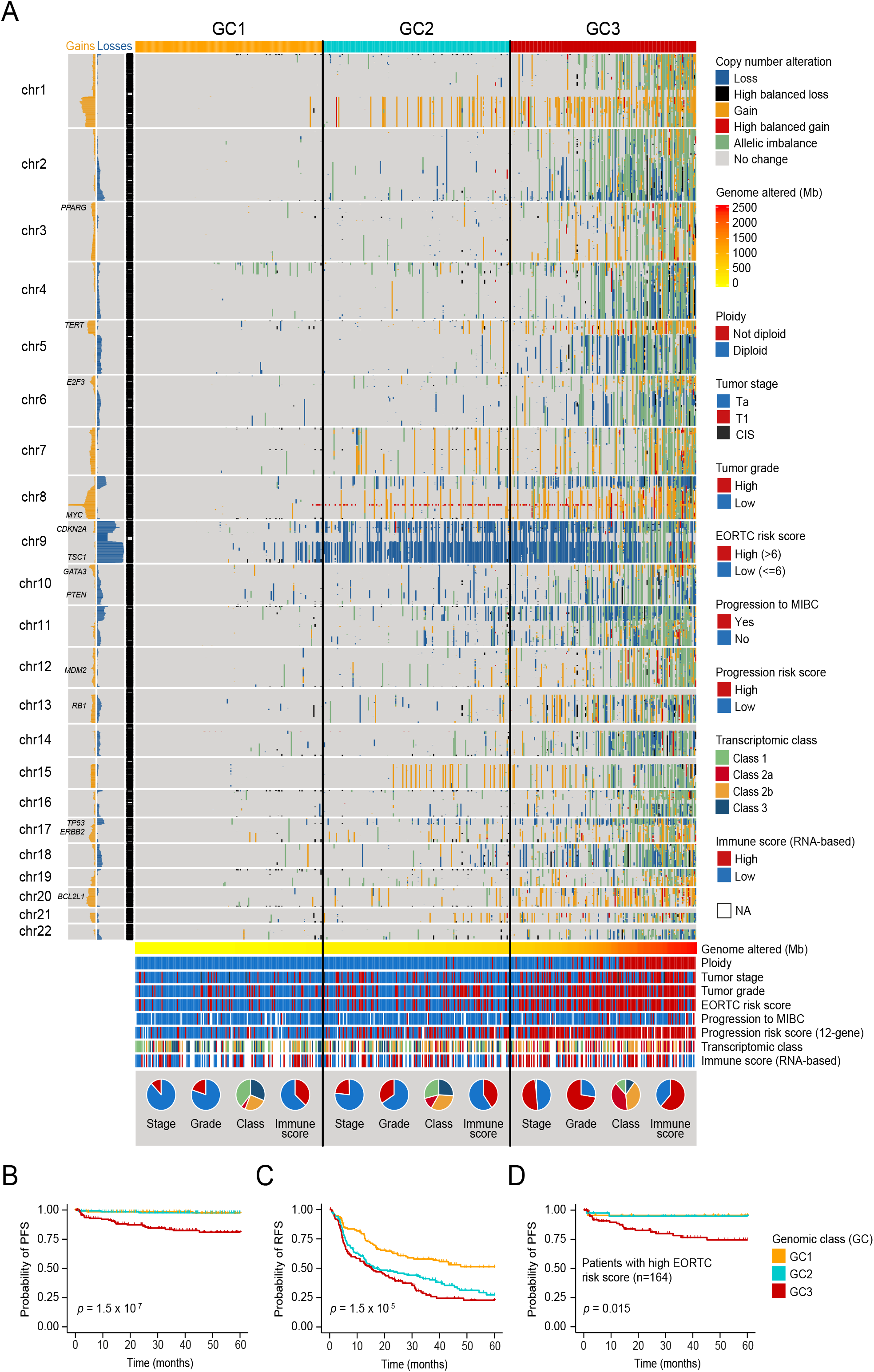
Copy number alterations in NMIBC. **a** Genome-wide copy number landscape of 473 tumors stratified by genomic class (GC) 1-3. Gains (gain + high balanced gain) and losses (loss + high balanced loss) are summarized to the left of the chromosome band panel. **b** Kaplan-Meier plot of PFS for 426 patients stratified by genomic class. **c** Kaplan-Meier plot of RFS for 399 patients stratified by genomic class. **d** Kaplan-Meier plot of PFS for patients with high EORTC risk score (n=164) stratified by genomic class.

### Integration of genomic alterations and transcriptomic classes

Integrative analysis of genomic and transcriptomic data from 303 tumors showed that transcriptomic classes were significantly associated with genomic classes (p=0.0005; **Fig 3A**). Class 2a included the highest fraction of tumors in GC3 (68%; 39/57). To document this association further, we found a strong correlation between genomic classes and progression risk score (n=449, p=3.24 × 10^−41^), and tumors with a higher progression score were predominantly class 2a and 2b (p=1.8 × 10^−32^; **Fig 3B**). When analyzing class 2a and 2b tumors only, genomic classes were still significantly associated with PFS; all progression events were associated with GC3 tumors (**Fig 3C**). Likewise, when analyzing genomically high-risk (GC3) tumors only, transcriptomic class 2a and 2b were still associated with PFS (p=0.036, **Fig S3A**).

**Figure 3.**
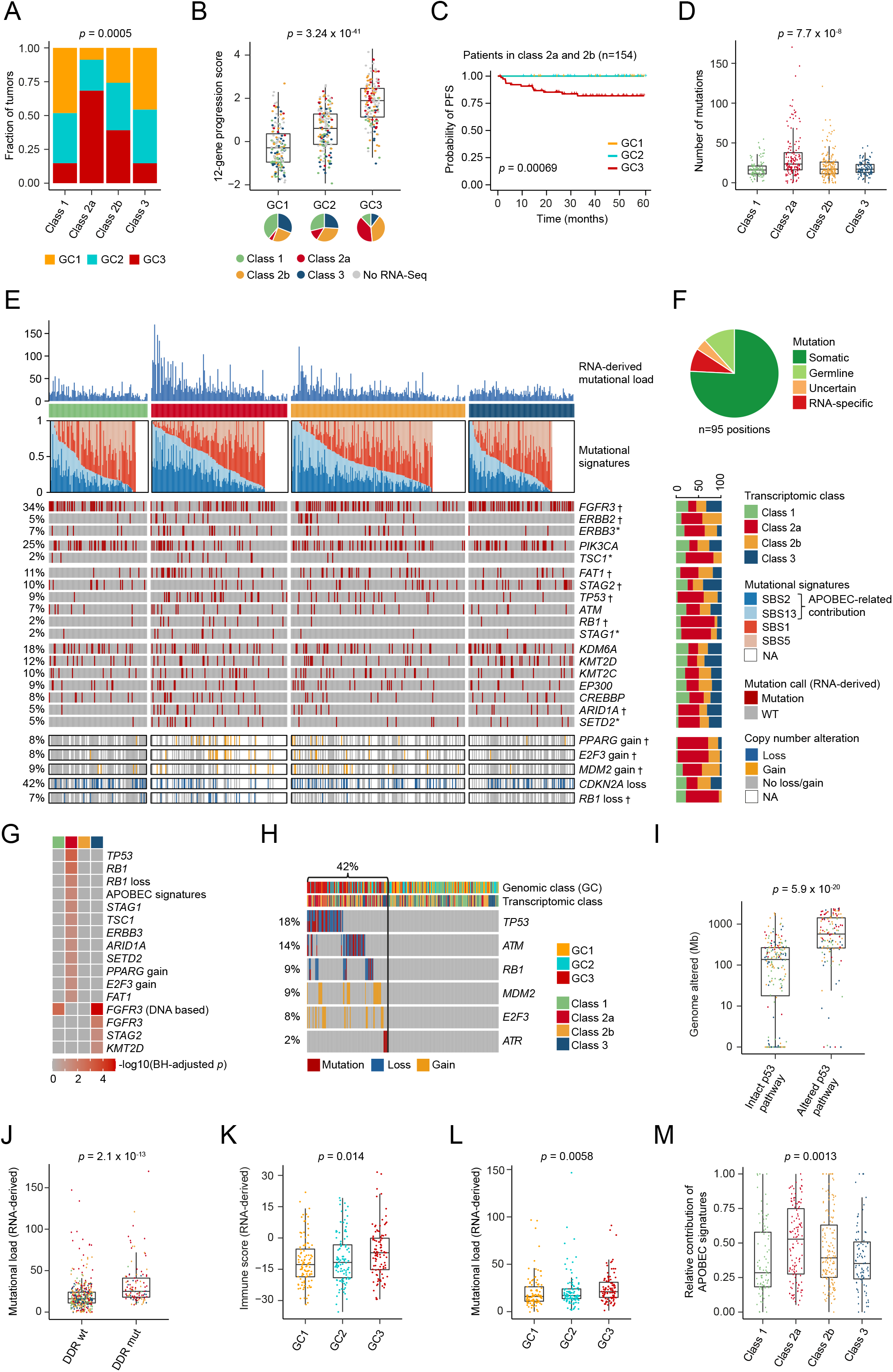
Genomic alterations associated with transcriptomic classes. **a** Genomic classes (GCs) compared to transcriptomic classes (n=303). **b** 12-gene qPCR-based progression risk score compared to GCs. Colors indicate transcriptomic classes. **c** Kaplan-Meier plot of PFS for 154 patients (including only class 2a and 2b tumors) stratified by GC. **d** Number of mutations according to transcriptomic classes. **e** Landscape of genomic alterations according to transcriptomic classes. Samples are ordered after the combined contribution of the APOBEC-related mutational signatures (SBS2+SBS13). Panels from the top: RNA-derived mutational load, relative contribution of four RNA-Seq-derived mutational signatures (inferred from 441 tumors having more than 100 single nucleotide variations), selected RNA-Seq-derived mutated genes, copy number alterations in selected disease driver genes (derived from SNP arrays). Asterisks (*) indicate *p* values below 0.05. Daggers (†) indicate BH-adjusted *p* values below 0.05. **f** Comparison of RNA-derived SNVs to whole-exome sequencing data from 38 patients, including 95 positions (non-hotspots) in the genes shown in E). We called four SNVs “uncertain” as they were observed in DNA but with few reads. **g** Genomic alterations significantly enriched in one transcriptomic class vs. all others. **h** Overview of p53 pathway alterations for all tumors with available copy number data and RNA-Seq data (n=303). **i** Amount of genome altered (Mb) according to p53 pathway alteration. **j** Number of mutations according to mutations in DNA-damage response (DDR) genes (including *TP53, ATM, BRCA1, ERCC2, ATR, MDC1*). **k** RNA-based immune score according to GCs. **l** Number of mutations according to GCs. **m** Relative contribution of the APOBEC-related mutational signatures (SBS2+SBS13) according to transcriptomic class.

Single-nucleotide variants (SNVs) with moderate or high functional impact were called based on RNA-Seq data. Class 2a tumors showed a significantly higher number of SNVs compared to all other classes (p=7.7 × 10^−8^; **Fig 3D**). Selected frequently mutated genes in bladder cancer are listed in **Fig 3E**, and a complete list of the most frequently mutated genes and genes with significantly different mutation patterns across classes can be found in **Fig S3B**. We compared RNA-derived SNVs (95 non-hotspot positions in the 18 genes shown in **Fig 3E**) with whole-exome sequencing of tumors from 38 patients, and validated 87% of the SNVs (73 somatic and 11 germline mutations) in the DNA (**Fig 3F**). Analysis of hotspot mutations in *FGFR3* (64%), *PIK3CA* (26%), *RAS* (7%), and *hTERT* (79%) based on tumor DNA is shown in **Fig S3B**. Furthermore, copy number alterations (from SNP microarray analysis) in disease driver genes are highlighted for comparison, and indicate overall loss of *CDKN2A*, significant gain of *PPARG* and *E2F3* in class 2a and loss of *RB1* in class 2a (**Fig 3E**). An overview of genomic alterations significantly associated with transcriptomic classes is shown in **Fig 3G**. Notably, p53 pathway alterations, observed in 42% of tumors (127/303; **Fig 3H**), were significantly associated with a high CNA burden (p=5.9 × 10^−20^; **Fig 3I**) and class 2a tumors (p=2.8 × 10^−7^). Gene expression levels of key molecules in the p53 pathway (*MDM2, E2F3, TP53, ATM* and *RB1*) were significantly correlated to the observed genomic changes (**Fig S3C-G**). Mutations in DNA damage repair (DDR) genes were significantly associated with RNA-derived mutational load (p=2.1 × 10^−13^; **Fig 3J**). This remained significant when *TP53* mutations were excluded from the analysis (p=4.4 × 10^−11^). In addition, we found a significantly higher mutational load and immune cell infiltration, estimated from RNA-Seq data, in GC3 tumors (**Fig 3K-L**), suggesting that these tumors present a high level of neoantigens.

Furthermore, we inferred seven trinucleotide single-base mutational signatures (**Fig S3H**), and four signatures showed high correlation to signatures previously identified in bladder cancer ^27,33,34^: SBS1 (age-related), SBS2 and SBS13 (related to excessive APOBEC activity) and SBS5 (related to *ERCC2* mutations; ^35^) (**Fig 3E**). Class 2a tumors showed significantly more mutations in the context of the APOBEC-related signatures (**Fig 3M**). Concordantly, high contribution of the APOBEC-related signatures was associated with worse PFS (**Fig S3I**), indicating that APOBEC activity may drive disease evolution and tumor aggressiveness^8^

Finally, we applied a deconvolution method (WISP; weighted in silico pathology) to assess intra-tumor heterogeneity and class stability from bulk transcriptomic profiles (Blum et al., 2019). WISP calculates pure population centroid profiles from the RNA-Seq data and estimates class weights for each sample based on the centroids (for details see **Methods**). We found that samples exhibited heterogeneity in all classes, with class 2a having the highest degree of heterogeneity and class 3 the lowest (**Fig S4A**). Associations of WISP class weights to molecular and clinical features were consistent with the previous description of the classes (**Fig S4B-D**). Class 1 weights were associated with lower tumor stage, tumor size and EORTC risk score. Class 2a weights were associated with *TP53* (p=4.51 × 10^−9^) and *TSC1* (p=1.37 × 10^−5^) mutations, as well as to higher tumor grade, tumor stage and EORTC risk score. Class 2b weights were significantly correlated to infiltration by all tested immune- and stromal populations (p<10^-10^). In addition, class 3 weights were associated with *FGFR3* and *PIK3CA* mutations, as well as lower tumor stage and grade. WISP class weights also outlined differences between class 1 and 3 signals: high class 1 weights were associated to *RAS* mutations and infiltration by myeloid dendritic cells, while high class 3 weights were strongly associated to *FGFR3* mutations (p=7.34 × 10^−15^) and lower immune and stromal population scores than the other classes.

### Spatial proteomics analysis of tumor and immune cell contexture

To resolve the immune features described above at the spatial level, multiplex immunofluorescence (mIF) and immunohistochemical (IHC) analyses were performed on 167 tumors, where additional tissue was available, targeting immune cells (T-helper cells, CTLs, Tregs, B-cells, M1- and M2 macrophages; see **Fig S5A** for details), carcinoma cells (pan-cytokeratin, CK5/6 and GATA3) and immune recognition/escape mechanisms (PD-L1 and MHC class I). Automated image analysis algorithms were developed to study the spatial organization of immune cells and immune evasion mechanisms (**Fig 4A** and **Fig S5A**). RNA-Seq data was available for 150 of the tumors and the RNA-derived immune score correlated significantly with the level of immune infiltration (p=3.3 × 10^−9^; **Fig 4B**). The different subsets of lymphocytes were predominantly present simultaneously in the stroma and the tumor parenchyma. Consequently, only a few tumors belonged to the immune excluded phenotype ^36^, and we therefore focused on the degree of infiltrating immune cells located in the tumor parenchyma, henceforth termed immune infiltration. Notably, tumors with a high immune infiltration showed a high expression of MHC class I (p=6.18 × 10^−12^). Only a few tumors expressed PD-L1 in the tumor parenchyma, and the majority of these tumors were highly inflamed (p=1.64 × 10^−5^; **Fig 4B**).

**Figure 4.**
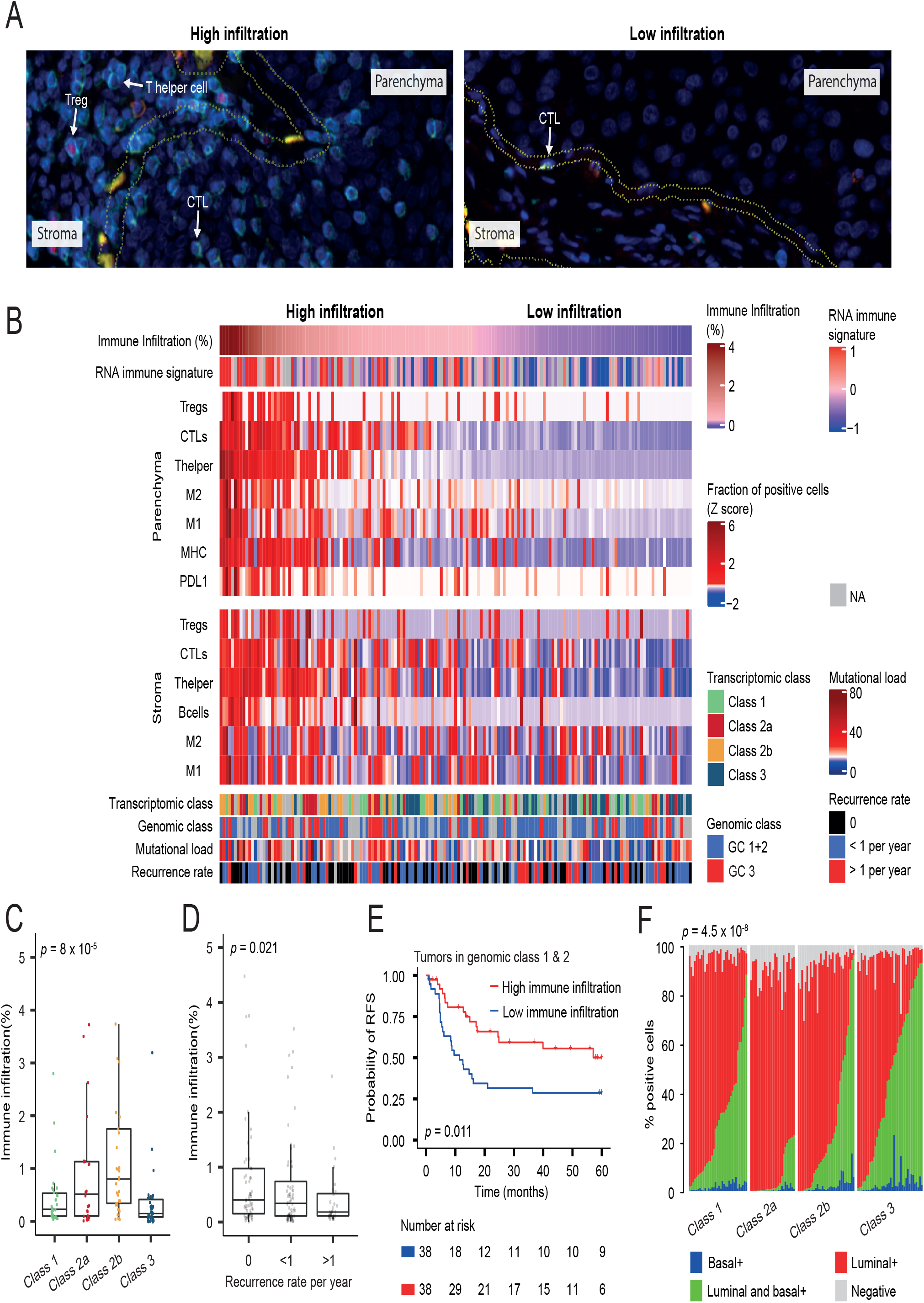
Spatial proteomics analysis of tumor immune contexture. **a** mIF staining with Panel 1 (CD3, CD8 and FOXP3) of a tumor with high- and low immune infiltration, respectively. **b** Spatial organization of immune cells and antigen recognition/escape mechanisms (MHC1 and PD-L1). The immune cells and immune evasion markers are defined as percent positive cells in the different regions (stroma and parenchyma) and normalized by z-scores. **c** Immune infiltration according to transcriptomic class. Immune infiltration is defined as the percentage of cells in the parenchyma classified as immune cells. **d** Immune infiltration stratified by recurrence rate. *p* value is calculated by the Jonckheere-Terpstra test for trend. **e** Kaplan-Meier plot of RFS stratified by immune infiltration in tumors with few genomic alterations (GC1+2). **f** Distribution of CK5/6 and GATA3 expression in carcinoma cells stratified by transcriptomic class. Each column represents a patient. The *p* value reflects the difference in CK5/6 expression across classes.

The level of infiltrating immune cells identified at the proteomic level was significantly associated with transcriptomic classes and class 2b tumors showed the highest immune infiltration (p=8 × 10^−5^; **Fig 4C**), supporting the observations delineated from the transcriptomic deconvolution analysis. These differences among transcriptomic classes were particularly evident for T helper cells and CTLs (p=2.5 × 10^−5^ and 0.0082, respectively; **Fig S5B**). These data confirm that the transcriptomic-based estimation of inflammation in class 2b tumors truly represents high immune cell infiltration.

Despite the overall aggressive characteristics of the inflamed class 2b tumors, a high immune infiltration was significantly associated with a lower recurrence rate (p=0.021; **Fig 4D**), particularly for T helper cells and CTLs (p=0.019 and 0.012, respectively; **Fig S5C**). There were too few progression events to document this effect on PFS. Furthermore, a possible protective immune response is shown in patients with tumors of similar genomic background (few genomic alterations); in this group, patients with high immune infiltration had a longer RFS compared to patients with low immune cell infiltration (p=0.011; **Fig 4E**).

Finally, we stained for basal cytokeratin expression (CK5/6) and luminal characteristics (GATA3) and aligned these with a pan-cytokeratin staining of the carcinoma cells to estimate the proportion of carcinoma cells positive for CK5/6, GATA3 or both (**Fig S5D**). All tumors stained positive for GATA3 and 23% expressed CK5/6 (>50% positive cells). All CK5/6 positive tumors were concurrently GATA3 positive and thereby not basal/squamous by definition ^37^. Similar coexpression of basal- and luminal-like markers has been observed previously in the Urothelial-like B tumors ^38^. The fraction of CK5/6 positive cells was strongly associated with transcriptomic classes, with class 3 having the strongest enrichment for CK5/6 expression (p=4.5 × 10^−8^; **Fig 4F**).

### Integrative prediction models, classifier construction and independent validation

An overview of the univariate Cox regression analyses of selected clinical features and molecular variables is shown in **Fig 5A**. In addition, we performed receiver operating characteristic (ROC) analysis for predicting progression using logistic regression models (n=301, **Fig 5B**). Combining EORTC risk score with genomic classes increased the predictive accuracy from 0.77 to 0.82, and combining EORTC risk score and transcriptomic classes increased the predictive accuracy to 0.85. Including all three variables in the model slightly increased the predictive accuracy to 0.87 (BH-adjusted p=0.036, Likelihood ratio test; full model vs. EORTC model). A logistic regression model of continuous variables (EORTC, genome altered and 12-gene progression score) showed no increased predictive value when including both molecular variables in the model (**Fig S6A**).

**Figure 5.**
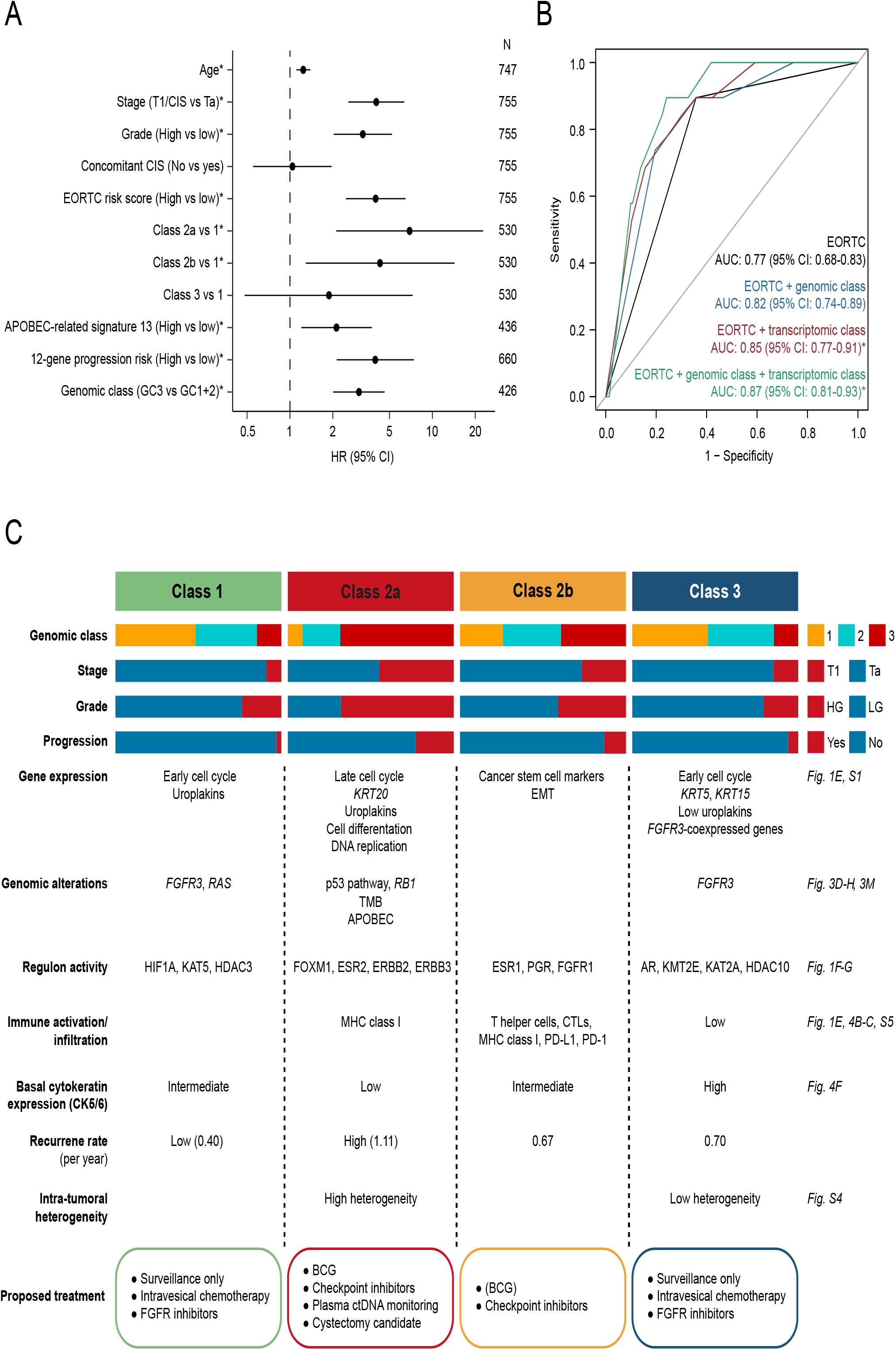
Prediction models and summary characteristics of transcriptomic classes. **a** Overview of hazard ratios calculated from univariate Cox regressions of PFS using clinical and molecular features. Black dots indicate hazard ratios and horizontal lines show 95% confidence intervals. *P* values below 0.05 are indicated by asterisks. **b** Receiver operating characteristic (ROC) curves for predicting progression using logistic regression models (n=301, events=19). Asterisks indicate significant model improvement compared to the EORTC model (Likelihood ratio test, BH-adjusted *p* value below 0.05). AUC=area under the curve. CI=confidence interval. **c** Summary characteristics of the transcriptomic classes. Molecular features associated with the classes are mentioned, and suggestions for therapeutic options with potential clinical benefit are listed. LG=Low grade, HG=High grade.

To facilitate the use of the four transcriptomic classes in future research and clinical settings, we constructed a single-sample classifier for NMIBC. The classifier was built similarly to the recently published tool for the consensus subtypes of MIBC. A class label is assigned to the transcriptomic profile of a tumor based on correlation to the class-specific mean expression profiles (^26^; for details see **Methods**). We applied the classifier to 15 independent cohorts, including five unpublished datasets, with a total of 1306 patients whose tumors were analyzed with a wide range of platforms (**Fig 6A**). Notably, RNA-Seq platforms were better suited to call class 3 tumors compared to microarray analyses. Overall, we found highly significant correlations between class and tumor stage, tumor grade and mutations in *FGFR3* and *TP53* (**Fig 6B**), and classes showed significantly different PFS (p=0.0002; **Fig 6C**) where patients with class 2a tumors had the worst outcome. Notably, multivariable Cox regression analysis revealed that class 2a (HR=2.9 (95%CI: 1.55-5.39); p=0.0009) and class 2b (HR=2.2 (95%CI: 1.03-4.46); p=0.041) were independently associated with worse PFS compared to class 1 when adjusted for tumor stage (**Table S6**). To validate the classifier further, we compared differences of regulon activity and biological pathway enrichment between classes in the discovery cohort to findings in the independent cohorts. The regulon and pathway analysis documented a high concordance between datasets (**Fig 6D-E** and **Fig S6B**), supporting the robustness of the classes.

**Figure 6.**
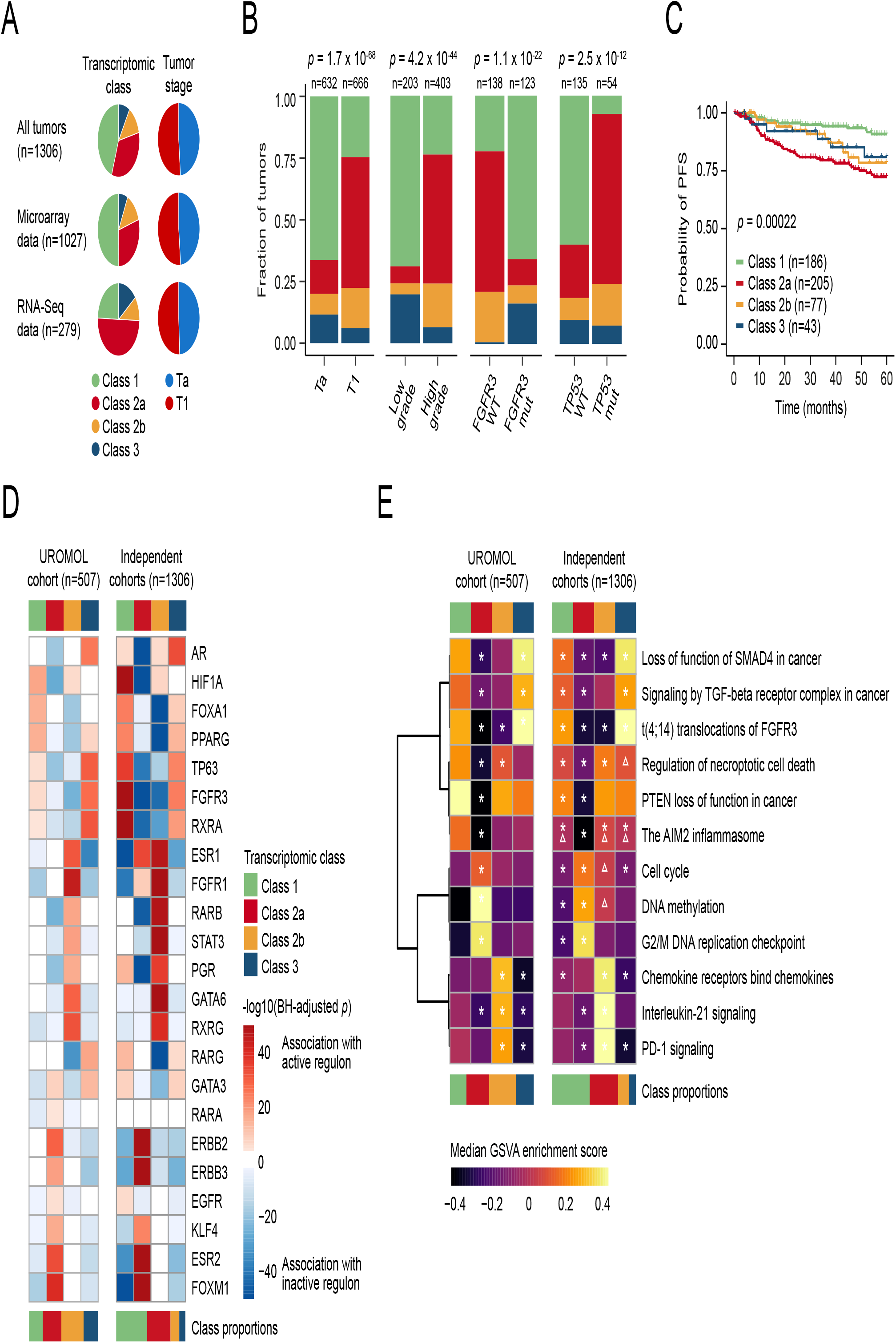
Validation of transcriptomic classes in independent cohorts. **a** Summary of classification results and stage distribution for all tumors, tumors with microarray data and tumors with RNA-Seq data. **b** Association of tumor stage, tumor grade and *FGFR3* and *TP53* mutation status with transcriptomic classes. **c** Kaplan-Meier plot of PFS for 511 patients stratified by transcriptomic class. **d** Association of regulon activities (active vs. repressed status) with transcriptomic classes in the UROMOL cohort (including samples with positive silhouette scores, n=507) and transcriptomic classes in the independent cohorts (pooled). The heatmap illustrates BH-adjusted p-values from Fisher’s Exact Tests. **e** Pathway enrichment scores within transcriptomic classes in the UROMOL cohort (including samples with positive silhouette scores, n=507) and transcriptomic classes in the independent cohorts (pooled). Asterisks indicate significant association between pathway and class (one class vs. all other classes, Mann-Whitney U-test, BH-adjusted p-value below 0.05). Triangles indicate direction swaps of pathway enrichment in the independent cohorts compared to the UROMOL cohort.

## Discussion

Here we expanded our analysis of NMIBC biology and associated clinical outcomes to 834 patient samples from the UROMOL consortium’s multicenter study. Utilising integrative multi-omics analysis, we demonstrated that disease aggressiveness in NMIBC patients was associated with genomic alterations, transcriptomic classes and immune cell infiltration. We described the development and validation of a single-sample transcriptomic classifier for NMIBC, and notably, identified patients with high chromosomal instability and poor outcome, denoted as class 2a. Importantly, we demonstrated that the genomic and transcriptomic subtypes showed strong independent prognostic value when compared to clinical risk factors. Integrative disease models of clinical risk factors and molecular features showed that the addition of transcriptomic class or genomic instability measures result in similar significant increases in AUCs, and the inclusion of both variables in disease models minimally improved the predictive accuracy (**Fig 5B** and **S6A**). Future classification schemes that incorporate all predictive molecular features may be optimal, however, for clinical application, we suggest the use of a transcriptomic-based classifier, as the expression data in addition will reflect tumor biological processes and possible treatment options (**Fig 5C**). The classifier was successfully validated using data from 1306 yet unpublished- and previously published patient samples.

Specifically, we showed that the extent of genomic alterations in NMIBC is an independent predictor of recurrence and progression. Tumors with high chromosomal instability should therefore optimally be managed as high-risk tumors regardless of histopathological findings. In addition, we demonstrated that the number of genomic alterations is significantly associated with high-risk transcriptomic classes, p53 pathway alterations and increased immune cell infiltration. Previous studies that applied array-based CGH analysis have shown that genomic alterations were correlated to histopathological parameters such as stage and grade ^13,14,39^, but independent prognostic value has not previously been described. We investigated a large clinically well-annotated patient series and applied SNP array technology to increase the granularity of genomic analysis and to gain information of allelic imbalance - a molecular feature not available through CGH analysis. It is noteworthy that, in the current study, we did not aim at identifying specific genomic loci associated with progression to MIBC, but instead we report that the overall CNA burden is directly associated with clinical outcome. This observation is in agreement with other findings linking chromosomal instability to intra-tumor heterogeneity, disease aggressiveness and poor patient outcome in various human tumor types ^40,41^. In bladder cancer, chromosomal instability has previously been linked to advanced muscle-invasive disease ^42^. Our observation is further strengthened by the identification of mutations in DDR genes and p53 pathway alterations which were associated with genomic instability. This link has been observed previously in a smaller set of bladder tumors ^42^. The underlying mechanisms responsible for the genomic instability is, however, not fully understood, but may be caused by oncogene activation and replication stress, which triggers DDR checkpoints ^43^. Mutations in DDR genes and p53 pathway alterations are therefore likely to cause the genomic instability observed. We used RNA-Seq data for mutation calling, which is associated with some limitations as only mutations in expressed genes can be detected and no germline reference is used to eliminate germline variations. However, we applied very stringent filters to avoid false positives and validated the presence of 87% of the RNA-Seq-derived mutations. Furthermore, a recent study also documented that high-precision analysis of mutations based on RNA is achievable ^44^.

At the transcriptomics level, we identified four main classes of NMIBC with the UROMOL2016 defined class 2 being separated into two new groups: class 2a and class 2b. Class 2a, displaying a higher RNA-derived mutational load and elevated APOBEC-related mutational signature contribution, was characterized as a high-risk group with multiple progression events, whereas class 2b, displaying higher expression of stem cell and EMT markers and immune infiltration, was associated with a lower risk of progression. APOBEC-associated mutations are proposed to drive tumor evolution and disease aggressiveness in lung cancer ^45,46^ and high levels of the APOBEC3B protein have been associated with poor outcome in breast cancer ^47^. High tumor mutational burden and APOBEC mutational load have, however, previously been associated with a better prognosis of MIBC ^27^. Possible reasons for this discrepancy could be related to better treatment efficacy for muscle-invasive tumors with a high mutational burden and APOBEC-related mutational signatures ^3^.

The ability to discriminate between class 1 and class 3 tumors was possible since we analyzed a very large number of samples by RNA-Seq, a technology with much higher resolution than microarrays used in prior studies. Methylation analysis further emphasized the distinctive features of these two classes (**Fig S1C-D**). We also observed that class 3 tumors showed a high level of keratin 5 gene expression and simultaneously the highest level of CK5/6 protein expression; however, this should not be associated with basal/squamous MIBC tumors, since we also observed GATA3 expression in all of these tumors (see example in **Fig S5D**). The transcriptomic classes were prognostic *per se*, which further highlights several aspects of tumor biology (**Fig 5C**). Since all MIBC tumors initially arise as NMIBC, a relevant question is whether the recently developed MIBC consensus classification ^26^ would be applicable to NMIBC. We provide strong evidence that this is not the case (**Fig 1D**). Our analysis shows that NMIBC displays less dramatic phenotypic variability compared to MIBC, and classifiers have to be adjusted accordingly. The NMIBC classes described here overlap partially with previously generated signatures of outcome and gene expression subtypes in NMIBC ^6,8,48^. The subtypes from the Lund group initially generated based on the whole spectrum of bladder tumors ^7^, have now been further developed to include five major tumor cell phenotypes ^25,49^. As the classification system spans a large biological range (NMIBC to MIBC), it may not fully capture the subtype granularity observed exclusively in NMIBC. In our work, we compared our transcriptomic classes to the Lund classes using the Lund single-sample classification system ^49^. Although we observed an overlap between e.g. class 2a and GU, most tumors were classified as UroA.

Our analysis of regulons revealed potential druggable pathways related to sex hormones in distinct tumor subsets: the androgen receptor pathway was significantly activated in class 3 tumors, although there was no enrichment for male patients in this group. In a recent study, low levels of the androgen receptor have been linked to increased translation and tumor proliferation in prostate cancer ^50^, and the high levels observed in class 3 could therefore have a protective effect. Class 2a was dominated by high levels of ESR2 regulon activity, while 2b was dominated by high levels of ESR1 and PGR, indicating that hormonal receptor activity may play a pivotal role in disease development. The estrogen and androgen receptors have been linked to urothelial tumorigenesis in animal models ^51–53^. Furthermore, the androgen receptor has been shown to be specifically expressed in early-stage bladder tumors ^54^, corroborating the finding of a unique class with androgen receptor activity in NMIBC. It is, however, important to emphasize that the transcriptomic analyses of regulon activity were based on bulk tumor analysis and some regulon activities could therefore be driven by different tissue compositions, e.g. higher immune infiltration in class 2b tumors.

The different biological characteristics of the transcriptomic classes suggest that specific therapeutic interventions may have different effects in these patients, as outlined in **Fig 5C**. Of note, class 2a tumors were characterized by a high RNA-derived mutational load, which is considered to result in an elevated neoantigen burden, and these patients may therefore benefit from immunotherapy. Checkpoint inhibitors have been shown to be most effective in tumors with high mutational burden ^30^. Class 2b tumors were frequently PD-L1 positive, suggesting that these patients may also benefit from checkpoint inhibitor immunotherapy, since high PD-L1 has been linked to an improved response to both PD-1 and PD-L1 inhibitors in MIBC ^55,56^. The interest towards the use of systemic immunotherapy in NMIBC has gained momentum, and Pembrolizumab (PD-1 inhibitor) has recently been approved by the FDA for BCG-unresponsive NMIBC patients. The frequent *FGFR3* mutations observed in class 1 and 3 suggest that FGFR inhibitors could be effective in these tumors, especially since the oral FGFR inhibitor BGJ398 recently showed antitumor activity in a marker lesion study of patients with NMIBC (^57^, NCT02657486), and Pemigatinib (FGFR1,2,3 inhibitor) is being tested in an ongoing phase II clinical trial in patients with recurrent low- or intermediate-risk tumors (NCT03914794). Intravesical chemotherapy should be considered especially for class 3 tumors, but possibly also for class 1 tumors although the recurrence rate is lower in these patients.

BCG response mechanisms have been studied intensely ^58^ and so far one of the most promising markers of BCG response is fluorescence *in situ* hybridization (FISH, Urovysion) analysis of chromosomal abnormalities ^59^. A recent study of resistance to BCG treatment showed a higher baseline tumor PD-L1 expression among patients non-responsive to BCG compared to patients responsive to BCG treatment ^60^, indicating that the pre-treatment tumor microenvironment may play a crucial role in BCG response mechanisms. Thus, class 2b tumors, with the highest PD-L1 expression, may respond poorly to BCG. In this study, we did not observe any tumor-centric biological variables that were associated with BCG treatment response. However, the number of patients that received >5 cycles of BCG in connection with the analyzed tumor was low, and larger studies of BCG response are needed to delineate response mechanisms.

In conclusion, we report the largest integrative multi-omics analysis of NMIBC tumors from a total of 834 patients included in the UROMOL project. We delineate biological processes associated with disease aggressiveness based on detailed, high-quality clinical data, and we provide and validate a classification tool for assigning transcriptomic class and associated progression risk to independent samples. Transcriptomic classification of disease biology provides an important framework for novel biomarker discovery in next-generation clinical trials to optimize the current clinical management of patients with NMIBC.

## Data Availability

Normalized RNA read counts (accession# to be included, submission in progress) and SNP microarray data (accession# to be included, submission in progress) are deposited at the European Bioinformatics Institute (EMBL-EBI) Array Express. Raw sequencing data is deposited at The European Genome-phenome Archive (EGA) under accession numbers EGAS00001001236 and EGASxx (submission in progress). There are restrictions to the availability of raw sequencing data deposited at EGA due to Danish legislation regarding sharing and processing of sensitive personal data. Data can be shared if the new research purposes proposed by the data importers are approved by the National Committee on Health Research Ethics in Denmark. Furthermore, data processor agreements and contracts need to be signed to fulfil European GDPR data sharing rules. The lead contact will accommodate reasonable requests. Processed (non-sensitive) data will be shared upon request without restrictions.

## Acknowledgements

SVL, FFP, and LD are supported by the following funding sources: Aarhus University, The Danish Cancer Biobank, The Health Research Foundation of Central Denmark Region, The Danish Cancer Society. NM and FXR are supported by Fondo de Investigaciones Sanitarias (FIS), Instituto de Salud Carlos III, Spain (#PI18/01347), Asociación Española Contra el Cáncer (AECC, # GB28012014). DJD is supported by the following funding sources: RSG 17-233–01-TBE from the American Cancer Society, the W.W. Smith Charitable Trust, the Pennsylvania Department of Health via Tobacco CURE Funds, the Ken and Bonnie Shockey Fund for Urologic Research and the Bladder Cancer Support Group at Penn State Health. JIW is supported by the Laurence M. Demers Career Development Professorship in Pathology and Medicine, Pennsylvania State University. JDR is supported by the Ken and Bonnie Shockey Fund for Urologic Research at Penn State Health. JM is funded by a SEED Award from the HOPE Foundation, the Department of Defense (W81XWH-18-0257), and the VHA (BX003692-01). We thank KK Cheng, Maurice P Zeegers, Nicholas D James, Naheema S Gordon, Ben Abbotts and Roland Arnold for work involved in generating the Birmingham RNA-Seq validation cohort.

## Author contributions

Conceptualization, N.M., F.X.R., S.V.L, F.F.P., P.L., and L.D.

Methodology, S.V.L, F.F.P., P.L., A.T., C.S.G., K.B.D., T.S., I.N., E.C., M.S., N.B., L.M.S.,

T.S. (pathology), N.M., F.X.R., and L.D.

Formal Analysis, S.V.L, F.F.P., P.L., A.T., C.S.G., and M.S.

Investigation, S.V.L, F.F.P., P.L., A.T., C.S.G., M.S., N.M., F.X.R., and L.D.

Resources, K.B.D., J.B.J., G.G.H., A.C.P., V.W., M.G., M.H., G.S., M.H., K.M., R.N., B.J., X.L., D.D., D.W., R.A., C.D.H., J.D.R, J.I.W., U.S., D.S., K.E.M. van K., T.M., J.J.M., D.J.D., R.T.B., M.K., T.S. (Belgrade), A.H., E.Z., P.M., N.M., F.X.R., and L.D.

Data Curation, S.V.L, F.F.P., and P.L.

Writing - Original Draft, S.V.L, F.F.P., P.L., and L.D. Writing - Review & Editing, all authors.

Supervision, L.D.

Project Administration L.D.

Funding Acquisition, S.V.L., F.F.P., J.D.R., J.I.W., D.J.D., N.M., F.X.R., and L.D.

## Competing interests

L.D. has sponsored research agreements with C2i-genomics, Natera, AstraZeneca and Ferring, and has an advisory/consulting role at Ferring.

J.B.J. has sponsored research agreements with Medac, Photocure ASA, Cephaid, Nucleix, Astellas and Ferring, and has an advisory board role at Olympus Europe, Cephaid and Ferring.

J.D.R. is involved in a sponsored scientific study or trial with Pacific Edge Biotechnologies, MDxHealth and Urogen Pharma, is a consultant for Urogen Pharma, and has an investment interest in American Kidney Stone Management.

J. J. M. is a consultant for Ferring, AstraZeneca, Janssen and participated in advisory boards for Foundation Medicine and Nucleix.

## Methods

### Patients and data in the UROMOL discovery cohort

Patients in the discovery cohort were included in the UROMOL project and followed according to national guidelines. Further details regarding samples, procedures and clinical follow-up are listed in ^8^. The study was approved by the Central Denmark Region Committees on Biomedical Research Ethics (#1994/2920; Skejby, Aalborg, Frederiksberg); the Danish National Committee on Health Research Ethics (#1906019), the ethics committee of the University Hospital Erlangen (#3755); the ethics committee of the Technical University of Munich (#2792/10); Medical Ethics Committee of Erasmus MC (MEC#168.922/1998/55; Rotterdam); the Uppsala Region Committee on Biomedical Research Ethics (#2008/252); the Ethical Committee of Faculty of Medicine, University of Belgrade (#440/VI-7); the Ethics Committee (CEIC) of Institut Municipal d’Assistència Sanitària/Hospital del Mar (2008/3296/I); the ethics committee of the University Hospital Jena (#4774-4/16).

RNA-Seq data from 438 tumors included in our previous work ^8^ was reanalyzed together with new RNA-Seq data from 97 tumors. See **File S1** for details. Based on the discovery samples, we created a “BCG cohort” of 55 patients who meet the following criteria: 1) indication of BCG treatment was high-grade disease, 2) the patient received a minimum of six BCG series and 3) BCG treatment was initiated within 12 months after TURB (hence, BCG was given in relation to the analyzed tumor). The BCG cohort was used to investigate response to BCG treatment using multiple features available from our datasets.

### DNA and RNA extraction

Procedures for nucleotide extraction from tumors and leukocytes are described in ^8^.

### Total RNA-Sequencing

Sequencing of total RNA was performed using ScriptSeq-v2 RNA-Seq Library Preparation Kit (Illumina) and KAPA RNA HyperPrep Kit with RiboErase HMR (Roche). RNA input was 500 ng for both kits.

### Gene expression quantification and normalization

We remapped and requantified all new and previously generated expression data. Salmon ^61^ was used to quantify the amount of each transcript using annotation from GRCh38. The R packages tximport and edgeR were used to summarize the expression at gene-level and normalize the data, respectively.

### Consensus Clustering

The expression matrix was filtered to only include transcripts with a median expression above zero. Genes were ranked based on median absolute deviation (MAD) across all samples and divided into subsets of the top -2000, -4000, -6000, -8000, -10,000, -12,000 MAD-ranked genes. Consensus clustering was performed on the different gene subsets using the R package ConsensusClusterPlus (settings: maxK=10, reps=1000, pItem=0.95, pFeature=1, clusterAlg=“hc”, distance=“pearson”). To identify the most representative samples within each cluster, silhouette scores were computed for all samples using the R package CancerSubtypes. A four-cluster solution based on the top-4000 MAD-ranked genes was chosen.

### Gene expression signatures

We extracted genes associated with cell cycle, keratins, uroplakins, cancer stem cells, epithelial-mesenchymal/mesenchymal-epithelial transition, and differentiation ^7,62,63^ and summarized each biological process as the mean expression of all marker genes associated with the given process. Gene expression signatures of bladder cancer have previously been reported, including a progression signature and CIS signature ^4,5,64,65^. All 535 samples in the RNA-Seq cohort were classified according to the three signatures using consensus clustering. Finally, we characterized the classes using gene signatures of potential relevance for different treatment strategies ^7,26,30,31,66–69^.

### RNA-based estimation of immune cell infiltration

As in Rosenthal et al. ^29^, we evaluated immune cell infiltration based on the expression of predefined gene lists for 14 different immune cell populations ^70^ (for CD4^+^ T cells: ^71^). A score for each cell type was calculated as the mean expression of all marker genes associated with the given cell type, and a total immune score was defined as the sum of all immune cell type scores.

### RNA-based mutation calling

Mutations were called from the RNA-seq data using the GATK pipeline. Briefly, STAR v2.7 was used to align the raw RNA reads to the hg38 human genome assembly and PICARD tools were used to mark duplicates. GATK tools, SplitNCigarReads, BaseRecalibrator and ApplyBQSR were applied in order to reformat some of the alignments that span introns and correct the base quality score. Finally, the HaplotypeCaller software was used to call variants. The resulting VCF files were annotated using SnpEff followed by filtration for possible impact on proteins. First, only SNVs annotated with a HIGH or MODERATE impact by SnpEff were included and SNVs in splice-site genomic locations were excluded. Second, mutations with an rs ID in dbSNP were excluded. Third, only mutations with a quality score above 100 and a Fisher Strand score (FS) below 30.0 were included. Finally, mutations called in ten or more samples were filtered out with the exception of known mutation hotspots (*FGFR3* and *PIK3CA*). Furthermore, we validated RNA-derived mutations in DNA for a subset of patients (n=38) where whole-exome sequencing data were available.

### RNA-based mutational signature analysis

To infer mutational signatures, we included mutations called within the gene sequence (HIGH, MODERATE, and LOW impact) and excluded mutations with rs ID together with mutations with a quality score below 100 or a Fisher Strand score (FS) above 30.0. Finally, mutations were included if they met the following criteria: 1) alternate allele frequency (AF) >0.15 and <0.60; 2) number of reads >20. Only samples with more than 100 SNVs were kept to infer the mutational signatures (n=441). We used non-negative matrix factorization to decompose the motifs matrix into seven signatures and their corresponding weights using the R package SomaticSignatures. The similarity between the seven inferred signatures and defined COSMIC signatures was examined using the R package MutationalPatterns.

### Copy number analysis

GSA Illumina SNP arrays (∼760k positions) were used on tumor DNA from 473 patients in order to assess copy number alterations. We previously applied the Infinium OncoArray-500K BeadChipGenotyping arrays for the paired germline samples and used this as reference. LogR Ratio (LRR) and B-allele-fraction (BAF) were corrected and normalized using the Genotyping module from GenomeStudio 2.0 (Illumina) within each array type and all positions uniquely found in both arrays were exported for further analysis (151,291 probes). The R package ASCAT was used for segmentation of the genome and we used the raw-segmented total copy number, the raw-segmented BAF data and various empiric thresholds (gains: > 0.08, high gains: > 0.16, loss: <-0.1, high loss: <-0.2, allelic imbalance (AI): <0.45) to identify five different types of CNAs: 1) losses associated with AI (i.e., associated with a deviation in BAF), 2) gains associated with AI, 3) high losses without AI, 4) high gains without AI and 5) AI without a change in total copy number. The applied thresholds were validated using histograms of LRR and in all diploid cases (83%), the peak for no change in copy number was within the thresholds defined for the gains/losses without deviation in BAF. Using these thresholds, subclonal events present in a minority of carcinoma cells will either not be called or instead be defined as regions with no copy number changes but with deviation in the BAF (due to the higher sensitivity of the BAF measurement). Therefore, defined gains/losses are clonal events or subclonal events present in the majority of carcinoma cells.

The amount of genome in a non-normal state was calculated using the thresholds above (referred to as the CNA burden). Tumors were assigned to three genomic classes (GC1-3) of equal size based on the CNA burden to illustrate low, intermediate and high chromosomal instability (cut-offs at the 33rd and 67th percentiles). Furthermore, based on LRR and BAF plots, we manually defined tumors as being diploid or not diploid.

### Methylation analysis

DNA methylation analysis was performed using DNA from 29 patients based on the UROMOL2016 classification with 10-11 samples from each class. After re-classification, we had 10 samples in class 1, 12 in class 2a/2b with a majority in class 2b and 6 in class 3. All tumors were selected to have a high silhouette score, and all were Ta high-grade tumors. We used 500 ng genomic DNA for bisulfite conversion followed by whole-genome amplification prior to hybridization to EPIC BeadChip (Illumina, San Diego, CA) overnight as described by the manufacturer and then scanned with the Illumina iSCAN system. Data was imported and processed using the RnBeads v2.2 R package pipeline. For the preprocessing of the data, the normalization method was set to ‘illumina’ and the background correction method to ‘methylumi.noob’.

### Regulon analysis

We reconstructed transcriptional regulatory networks (regulons) consisting of 23 transcription factors and associated induced/repressed targets using the R package RTN, as previously described ^27^. Additionally, we investigated 78 candidate regulators associated with chromatin remodeling in cancer ^28^. Potential associations between a regulator and all possible target genes were inferred from the expression matrix by Mutual Information and Spearman’s correlation, and permutation analysis was used to remove associations with a BH-adjusted p-value > 1 × 10^−5^. Unstable associations were eliminated by bootstrap analysis (1000 resamplings, consensus bootstrap >95%) and the weakest association in triangles of two regulators and common target genes were removed by data processing inequality (DPI) filtering (tolerance=0.01). Regulon activity scores for all samples were calculated by two-tailed gene set enrichment analysis.

### 12-gene progression score

All molecular data related to the 12-gene progression score were generated previously ^6^ and analyzed here with additional follow-up information.

### Construction of single-sample transcriptomic classifier

We constructed a Pearson nearest-centroid classifier for NMIBC based on the recently published classifier for the MIBC consensus subtypes ^26^. Only samples with positive silhouette scores were used for feature selection (n=507). We filtered the expression matrix to include genes with a median expression > 0 in at least one of the four classes and used a step-wise ANOVA approach to identify genes with significantly different expression levels across classes. ANOVA between all four classes resulted in 13,650 significant genes (BH-adjusted p-values < 0.05). Genes highly expressed in class 2b dominated the list, so we removed class 2b samples and previously significant genes from the dataset and performed a second round of ANOVA on the remaining classes. This analysis added only four significant genes to the feature list (BH-adjusted p-values < 0.05). Next, class 2a samples were removed and one last round of ANOVA between class 1 and class 3 was performed (corresponding to a *t*-test), resulting in 109 significant genes (BH-adjusted p-values < 0.05). Thereby, a total number of 13,762 genes were suggested to be differentially expressed between classes. The step-wise ANOVA approach was chosen instead of multiple pairwise *t*-tests to reduce the number of statistical tests while still accessing differences between all classes. We computed the area under the curve (AUC) associated with each gene for prediction of the four classes and kept genes with an AUC > 0.6 (n=10,149). An additional filtering of genes was performed to only keep genes with a mean expression > 0 across all samples. Overall, the initial selection of features resulted in a list of 9,451 genes.

We used Leave One Out Cross-Validation (LOOCV) to assess the classification performance associated with different subsets of the 9,451 features. In each LOOCV run, we computed the mean fold-change associated with each gene for each class versus the others. Genes were ordered by their mean fold-change within each class and the four-gene lists were used to generate several gene subsets. The N top up-regulated and N top down-regulated genes within each class, with N varying from 50 to 800, were selected and used as feature input for the classifier. We obtained the lowest LOOCV error rate when selecting the 368 top up-regulated and 368 top down-regulated genes within each class (1,964 unique genes in total). Finally, genes appearing in >80% of the LOOCV runs were selected and used to build the final classifier (n=1,942). We computed four centroids corresponding to the four NMIBC classes (i.e. the mean gene expression profile of the 1,942 chosen feature genes for each class), and class labels are then assigned to single NMIBC samples based on the Pearson correlation between a sample’s expression profile and the four-class centroids. The NMIBC classifier is available as a web application at http://cit.ligue-cancer.net:3838/apps/BLCAclassify/.

### Proteomics

Formalin-fixed paraffin-embedded (FFPE) tissue from transurethral resection of bladder tumors (TURBT) was obtained from 167 Danish patients at Skejby and Frederiksberg hospital. Tissue microarrays (TMAs) were constructed from representative tumor areas with 1 mm triplicate core biopsies using the automated TMA-GRAND Master (3DHISTECH Ltd, Budapest, Hungary). Multiplex immunofluorescence analysis (mIF) was performed on TMA sections (3 μM) applying two panels of antibodies targeting CD3, CD8 and FOXP3 in the first panel and CD20, CD68, CD163 and HLA-A, B, C in the second (see **Table S5** for a detailed description of the panels). We utilized a tyramide signal amplification strategy on the BenchMark ULTRA staining instrument (Ventana) according to the manufacturer’s operating instructions. The fluorophore-labeled sections were imaged using the NanoZoomer s60 scanner (Hamamatsu). Immunostaining for pan-cytokeratin (Clone A1/A3, 1:100; Dako) as a second layer was performed on all mIF stained sections to identify carcinoma cells. For digital pathology, we utilized the Visiopharm image analysis software. For each tissue core, the pan-cytokeratin stained image was aligned to its corresponding fluorescence image. Intratumoral and stromal regions were defined automatically as pan-cytokeratin positive and negative, respectively. All immune cell populations from each panel were automatically characterized and quantified - and verified by an experienced pathologist (**Fig S5A**).

Identification of PD-L1 expression on the carcinoma cells was performed using two sequential TMA sections, the first section stained for pan-cytokeratin (Clone A1/A3, 1:100; Dako) and the second against PD-L1 (Clone Sp263, ready to use; Ventana). These sections were aligned and analyzed in a similar fashion as for the immune cell markers. Identification of basal and luminal markers on the carcinoma cells was performed using three sequential TMA sections, stained for pan-cytokeratin (Clone A1/A3, 1:100; Dako), GATA3 (Clone L50-823, ready to use; Ventana) and CK5/6 (Clone D5/16 B4, ready to use; Agilent/Dako) (**Fig S5D**). These sections were aligned and the proportion of carcinoma cells positive for GATA3, CK5/6 or double-positive was quantified for each tumor. Tumors were classified as positive if more than 50% of the carcinoma cells expressed the marker.

### Independent transcriptomics datasets used for validation

Transcriptomics data from ten historical cohorts (Kim ^72^, GEO: GSE13507; Lindgren ^39^, GEO: GSE32549; Sjödahl2012 ^7^, GEO: GSE32894; CIT ^66^, ArrayExpress: E-MTAB-1803; Choi ^73^, GEO: GSE48075; Sjödahl2017 ^25^, GEO: GSE83586; Song ^74^, GEO: GSE120736; Sjödahl2019 ^75^, GEO: GSE128959; Thorsen ^76^; Aaboe ^77^) were used for the validation of the four-class NMIBC classification. The data was downloaded from GEO or ArrayExpress and annotated with HUGO Gene Symbols. In addition to using the publicly available data, we included data from five yet unpublished cohorts (listed below). Each sample in each cohort was classified using the single-sample classifier trained using the UROMOL cohort.

*Unpublished cohort 1 provided by David DeGraff, Joshua Warrick and Jay Raman*: total RNA-Seq was obtained on 81 Ta tumors, all from formalin-fixed paraffin-embedded tissue, macro-dissected to enrich for tumor. Patients and materials from the Pennsylvania State University were retrospectively included and analyzed following Institutional Review Board approval and waiver of informed consent. RNA was extracted with Qiagen kits (Venlo, Netherlands). Sequencing was performed on the Illumina Novaseq 6000 instrument and run for 2×50 cycles. Only cases with alignment rate >85% were included. Expression was estimated from read counts using FPKM, then log2 transformed.

*Unpublished cohort 2 provided by Margaret Knowles*: Data from Affymetrix Human Transcriptome 2.0 microarrays for 104 stage T1 and 113 stage Ta tumors from the Leeds Multidisciplinary Research Tissue Bank (REC reference: 10/H1306/7). Total RNA was isolated from frozen tissue sections using a RNeasy Plus Micro Kit and amplified using the Affymetrix GeneChip WT PLUS Reagent Kit. The resulting cDNA was hybridised onto Affymetrix Human Transcriptome 2.0 microarrays. Quality control checks, gene level normalisation (using SST-RMA) and signal summarisation was conducted using Affymetrix Expression Console Software.

*Unpublished cohort 3 provided by Joshua Meeks*: RNA-seq based analysis of 73 FFPE tumors from T1 tumors treated with BCG at Northwestern University. All tumors were retrospectively included and analyzed following Institutional Review Board Approval and waiver of informed consent (STU00204352). RNA libraries were prepared using the Illumina TruSeq Stranded Total RNA Library Preparation Kit including rRNA depletion with RiboZero Gold. Libraries were sequenced on an lllumina HiSeq 4000, generating single-end, 50 bp reads. Trimmed reads were mapped to the GRCh37/hg19 genome with STAR v2.5.2. BAM files were processed to per-gene FPKMs with cuffquant from Cufflinks v2.2.1, using settings -u -p 12--library-type fr-firststrand, with an Ensembl v97 GRCh38 gene annotation GTF file, and cuffnorm, using default settings.

*Unpublished cohort 4 provided by Richard Bryan*: RNA-Seq based analysis of 85 tumors from the West Midlands Bladder Cancer Prognosis Programme (BCPP, ethics approval 06/MRE04/65), as previously described ^78^. RNA libraries were prepared using the Truseq Stranded Total RNA with Ribo-zero Gold kit (Illumina) and 2 × 100 bp PE sequenced (Hiseq, n=26) or 2 × 75 bp PE sequenced (Nextseq, n=52). The data were aligned to GRCh37 and reads counted with STAR aligner (v2.5.2b). Log2(Read count+1) for each gene has been used as input for the class prediction.

*Unpublished cohort 5 provided by Trine Strandgaard*: RNA-Seq based analysis of 47 fresh frozen tumors from patients enrolled at Aarhus University Hospital with high-risk NMIBC, and analyzed following approval by the the Danish National Committee on Health Research Ethics (#1708266). RNA-Seq data was generated using analysis pipelines described above for the additional samples included in the discovery cohort in this work.

### Pathway enrichment analysis

Pathway enrichment analysis was performed independently in the UROMOL cohort and each historical cohort that contained representatives of all classes. First, we collected pathway annotation from the Reactome (using R package reactome.db v1.68.0) and KEGG (using the R package KEGGREST v1.24.1) databases. We joined these annotations and performed gene-set variation analyses (using the R package GSVA v1.32.0) to obtain single-sample enrichment scores for each pathway.

To find associations between pathways and classes, we performed Mann-Whitney U-tests using the pathway enrichment scores between samples in each class versus samples in other classes in each cohort separately. *P* values were BH-adjusted. For the pathway visualizations, we first filtered pathways that were enriched in the same class in the UROMOL cohort and in at least 4 other datasets and then manually selected pathways from the filtered list. Pathway enrichment scores were grouped using hierarchical clustering with correlation distances (1 – *r*) and Ward clustering using the enrichment scores in the UROMOL cohort and the same pathway order was then used for the independent cohorts.

### Regulon activity in other cohorts

The regulons from the transcriptional networks calculated from UROMOL data were used to derive differential enrichment scores in each cohort separately using the two-tail GSEA method (R package RTNsurvival v1.8.3). We discretized the activity scores into ‘active’ and ‘repressed’ status, aggregated the regulon status in all cohorts, and used Fisher’s Exact Tests to find the association of regulon status with each class. *P* values were BH-adjusted.

### Weighted in Silico Pathology (WISP) analysis

To approximate intra-tumor heterogeneity, we used the bulk transcriptomic profiles and the consensus clustering results for the UROMOL cohort and applied the Weighted in Silico Pathology (WISP, R package v. 2.3) method with default settings. Only samples with a positive silhouette score were used for the WISP analysis (n=507). WISP consists of two main steps: 1) Calculation of pure population centroid profiles and 2) Estimation of pure population weights for each sample. First, WISP selects features for each class by iteratively considering ANOVA *p* values (FDR adjusted *p* values < 0.05), AUC scores (AUC > 0.8) and expression log-fold changes between classes, fitting a non-negative least squares model and removing samples considered mixed. A model is then built from the core of pure samples for each class (154 samples were kept as “pure” and 199 top marker genes were included in the centroid profiles). Next, WISP class weights were estimated for all the samples in the cohort (n=507) using the centroid profiles (hence, each sample is weighted between all four transcriptomic classes). We recovered the estimated WISP class weights and used Pearson and Spearman correlations to investigate their association to silhouette scores and MCPcounter immune scores, respectively. Finally, we used Mann-Whitney U-tests to associate WISP class weights to genetic mutations and clinical variables.

### Quantification and statistical analysis

Statistical comparisons between groups were performed using the Wilcoxon rank-sum test (Mann–Whitney U test) or Kruskal–Wallis test for continuous variables and Fisher’s Exact test, with Monte-Carlo simulations when necessary, for categorical variables. It is stated in the figure legends if tests other than the above-mentioned were applied. Survival analyses were performed using the Kaplan-Meier method and log-rank tests were used to compare survival curves (R packages survival and survminer). Cox Proportional-Hazards analyses were also performed using the R packages survival and survminer. We built logistic regression models to predict progression and used the predicted probabilities as variables in ROC analyses (R packages glmnet and pROC). AUCs and associated 95% CIs (computed with 2,000 stratified bootstrap replicates) were calculated using the R package pROC. Likelihood ratio tests were used to assess model improvement (all models were compared to the EORTC model). *P* values below 0.05 were considered significant across all tests and BH-adjustment of *p* values was performed to control for multiple testing when necessary (otherwise unadjusted *p* values are reported). All statistical and bioinformatics analyses were performed with R (v3.6.0 or 3.6.1).

## Data availability

Normalized RNA read counts (*accession# to be included, submission in progress*) and SNP microarray data (*accession# to be included, submission in progress*) are deposited at the European Bioinformatics Institute (EMBL-EBI) Array Express. Raw sequencing data is deposited at The European Genome-phenome Archive (EGA) under accession numbers EGAS00001001236 and EGASxx (*submission in progress*). There are restrictions to the availability of raw sequencing data deposited at EGA due to Danish legislation regarding sharing and processing of sensitive personal data. Data can be shared if the new research purposes proposed by the data importers are approved by the National Committee on Health Research Ethics in Denmark. Furthermore, data processor agreements and contracts need to be signed to fulfil European GDPR data sharing rules. The lead contact will accommodate reasonable requests. Processed (non-sensitive) data will be shared upon request without restrictions.

## Notes

### Funding Statement

Funding: SVL, FFP, and LD are supported by the following funding sources: Aarhus University, The Danish Cancer Biobank, The Health Research Foundation of Central Denmark Region, The Danish Cancer Society. NM and FXR are supported by Fondo de Investigaciones Sanitarias (FIS), Instituto de Salud Carlos III, Spain (#PI18/01347), Asociacion Espanola Contra el Cancer (AECC, # GB28012014). DJD is supported by the following funding sources: RSG 17 233 01 TBE from the American Cancer Society, the W.W. Smith Charitable Trust, the Pennsylvania Department of Health via Tobacco CURE Funds, the Ken and Bonnie Shockey Fund for Urologic Research and the Bladder Cancer Support Group at Penn State Health. JIW is supported by the Laurence M. Demers Career Development Professorship in Pathology and Medicine, Pennsylvania State University. JDR is supported by the Ken and Bonnie Shockey Fund for Urologic Research at Penn State Health. JM is funded by a SEED Award from the HOPE Foundation, the Department of Defense (W81XWH-18-0257), and the VHA (BX003692-01).
Ethichal review boards: The study was approved by the Central Denmark Region Committees on Biomedical Research Ethics (#1994/2920; Skejby, Aalborg, Frederiksberg); the Danish National Committee on Health Research Ethics (#1906019), the ethics committee of the University Hospital Erlangen (#3755); the ethics committee of the Technical University of Munich (#2792/10); Medical Ethics Committee of Erasmus MC (MEC#168.922/1998/55; Rotterdam); the Uppsala Region Committee on Biomedical Research Ethics (#2008/252); the Ethical Committee of Faculty of Medicine, University of Belgrade (#440/VI-7); the Ethics Committee (CEIC) of Institut Municipal dAssistencia Sanitaria/Hospital del Mar (2008/3296/I); the ethics committee of the University Hospital Jena (#4774-4/16).

